# Using compartmental models to simulate directed acyclic graphs to explore competing causal mechanisms underlying epidemiological study data

**DOI:** 10.1101/19007922

**Authors:** Joshua Havumaki, Marisa C. Eisenberg

## Abstract

Accurately estimating the effect of an exposure on an outcome requires understanding how variables relevant to a study question are causally related to each other. Directed acyclic graphs (DAGs) are used in epidemiology to understand causal processes and determine appropriate statistical approaches to obtain unbiased measures of effect. Compartmental models (CMs) are also used to represent different causal mechanisms, by depicting flows between disease states on the population level. In this paper, we extend a mapping between DAGs and CMs to show how DAG–derived CMs can be used to compare competing causal mechanisms by simulating epidemiological studies and conducting statistical analyses on the simulated data. Through this framework, we can evaluate how robust simulated epidemiological study results are to different biases in study design and underlying causal mechanisms. As a case study, we simulated a longitudinal cohort study to examine the obesity paradox: the apparent protective effect of obesity on mortality among diabetic ever-smokers, but not among diabetic never-smokers. Our simulations illustrate how study design bias (e.g., reverse causation), can lead to the obesity paradox. Ultimately, we show the utility of transforming DAGs into in silico laboratories within which researchers can systematically evaluate bias, and inform analyses and study design.

## 3 Introduction

Designing analyses to accurately estimate the effect of an exposure on outcome requires understanding how variables relevant to a study question are causally related to each other. Directed acyclic graphs (DAGs) are diagrams used in epidemiology to graphically map causes and effects to separate associations due to causality versus those due to bias. Compartmental model (CMs) depict parameterized flows between disease states over time [1, 2] and can be used to represent mechanisms, i.e. the explicit state transitions or processes sufficient for an exposure to lead to an outcome, underlying disease progression or transmission [3, 4]. CMs are often implemented using ordinary differential equations (ODEs), commonly linear ODEs with each variable representing the number of individuals in a distinct state (e.g. healthy vs. diseased). Compartmental models have a long history of use in medicine and public health see [5, 6] and references in [7]. Further details on the compartmental modeling framework are given in Appendix Section A1.1.

Given the causal nature of both DAGs and CMs, a question arises of whether these two approaches may be linked. Indeed, Ackley et al. provided a formal mapping from the basic building blocks of DAGs (e.g. causality, confounding and selection bias) to CMs [1]. See Figure 1 for an example illustration and Appendix Section A1.2 for a review and more in-depth comparison between DAGs and CMs. Using this mapping, a DAG and CM are defined as ‘corresponding’ if they represent the same conditional independencies. This correspondence represents an exciting new development in linking DAGs and CMs—here we expand this idea to a general framework for study design and sensitivity analysis in practice. This step is necessary to understand how simulating DAGs can provide actionable insight from the relationships between variables in a study and ultimately, inform study design and analyses. Additionally, designing this framework has allowed us to identify and develop approaches to handle practical problems encountered when translating real DAGs into CMs (e.g. combinatorial explosion).

**Figure 1:**
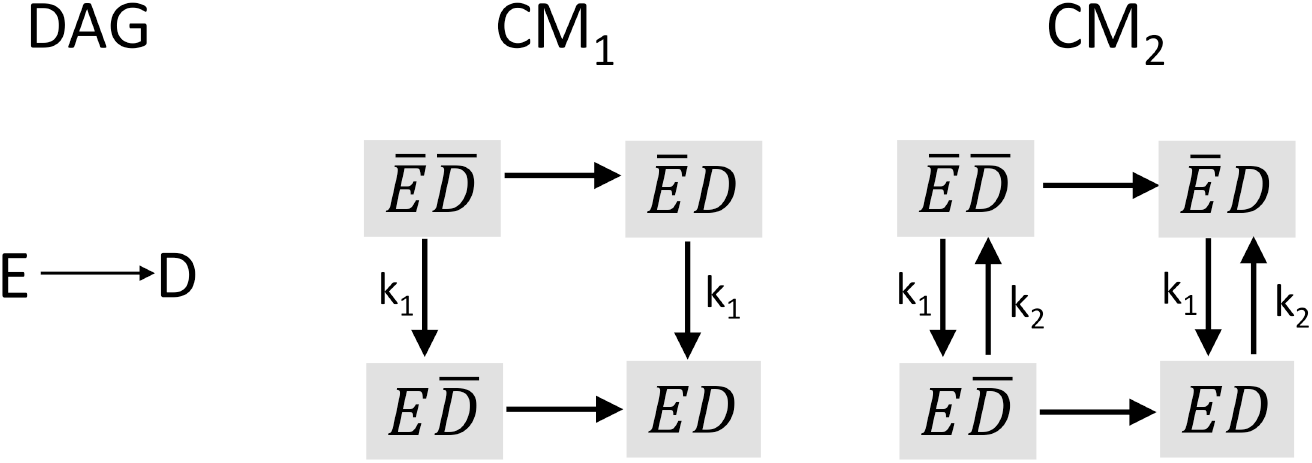
From left to right: A simple DAG showing causality wherein exposure *E* causes outcome *D*. Next, in *CM*_1_, We will assume that *E* and *D* are both dichotomous, so the corresponding CM will have 2*^n^* states (where *n* = 2 since there are 2 random variables on the DAG). Additionally, *D* status does not affect *E* status. The 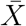 notation denotes *not* X, so *E* is unexposed. Thus the rates at which individuals become exposed (i.e. go from *Ē* to *E*) are the same whether or not they have *D*—we note that equal rates are denoted by the same parameter value and if the parameter symbol is not indicated, distinct rates are assumed, i.e. causal effects correspond to unmarked transition rates (in this example, the 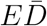 and 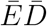 compartments have distinct transition rates for acquiring *D*, indicating a causal effect of *E* on *D*). This CM is further asserting that once an individual becomes diseased or exposed, they cannot return to the non-diseased or non-exposed state. In *CM*_2_, we see that individuals can move from *E* to *Ē*, but their *D* status does not affect the rate at which they transition as indicated by the equal rates *k_2_* between 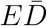 to 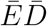 and *ED* to *ĒD*. Both *CM*_1_ and *CM*_2_ would be considered corresponding with the given DAG.

In this paper, we extended the work by Ackley et al. by developing an operationalized workflow which uses the mapping (between DAGs and CMs) to simulate epidemiological studies. We also note some opportunities to simplify this mapping to reduce the combinatorial explosion of CM compartments that results from realistic DAGs (taking advantage of simplifications to the CM that can be included when conditioning on a variable to reflect the make-up of the study population). We illustrated our findings by deriving a CM from a published DAG representing an instance of the obesity paradox, wherein obese ever-smoking diabetics have lower mortality rates than their normal weight counterparts. We examined competing hypotheses underlying the obesity paradox by incorporating different potential biases into our CM and then simulating study data. Our method can be applied to nearly any DAG or study question to gain insight into what underlying causal mechanisms can explain patterns observed in epidemiological data. This insight can be used to reduce bias in study designs and ultimately obtain more accurate effect measures of an exposure on outcome.

## 4 Methods

### 4.1 Overview of the Obesity Paradox

The obesity paradox is the apparent protective effect of obesity on mortality among individuals with chronic diseases such as heart failure, stroke, or diabetes [8–11].In this analysis, we used an observational study conducted by Preston et al. in which obese, ever-smoking (but not never-smoking) diabetics had lower mortality rates than their normal weight counterparts [9] as inspiration for our simulation study, because it is a clear example of an occurrence of the obesity paradox. We did not use the same dataset or aim to replicate the analysis or results in this study, rather we extended the published DAG representing the causal processes of interest, and used statistical analyses in the study to motivate examples of competing causal mechanisms that might be investigated. Figure 2(a) shows the published DAG from the observational study [9] representing the obesity paradox. The exposure is body mass index (BMI) and is coded as either overweight/obese (BMI ≥ 25 *kg/m*^2^) or normal weight (BMI = 18.5-24.9 *kg/m^2^*) and the outcome is mortality. Individuals are considered to have diabetes or prediabetes if their hemoglobin A1c is greater than 5.7%, or if they have been previously diagnosed. Smoking is a common risk factor for diabetes, mortality, and BMI, and is coded as ever-smoking (≥ 100 cigarettes over the course of an individual’s lifetime) or never-smoking (< 100 cigarettes). The mortality rates were age-standardized according to the 2000 census using age groups 40-59 and 60-74. For simplicity of notation, we will refer to prediabetics and diabetics as ‘diabetics’ and overweight and obese as ‘obese’.

**Figure 2:**
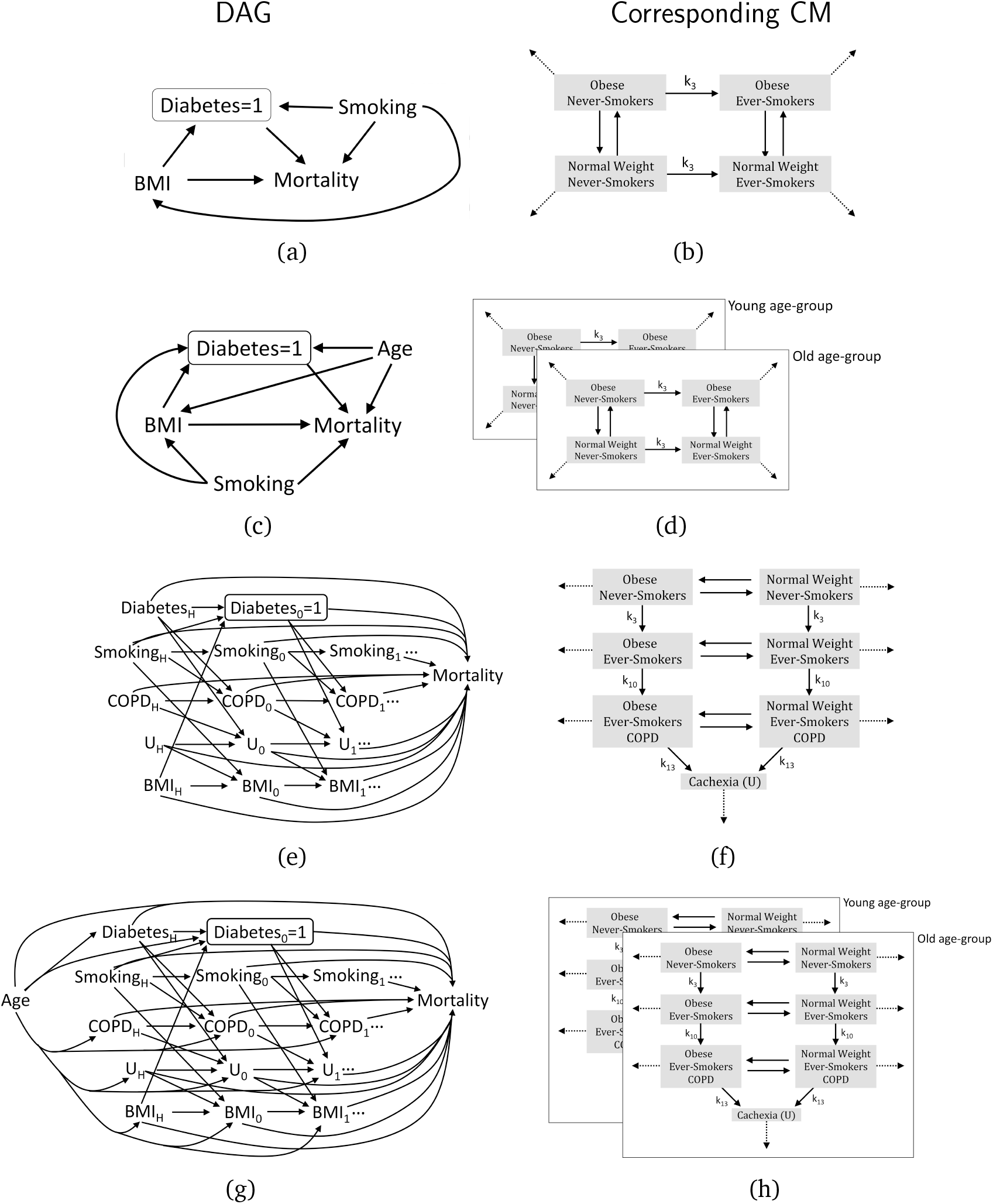
All DAGs and corresponding CMs used in our obesity paradox simulation study. DAGs (left column) and corresponding CMs (right column) for each model. (a) Preston et al. [9] DAG and (b) corresponding CM; (c) adding in age-varying mortality rates and (d) corresponding CM; (e) reverse causation and (f) corresponding CM; finally, (g) combined model and (h) corresponding CM. In the DAGs, ‘BMI’ is the exposure and ‘Mortality’ is the outcome. ‘Age’ and ‘Smoking’ confound the relation between ‘BMI’ and ‘Mortality’. The box around ‘Diabetes’ indicates that the study population is conditioned on a single strata of diabetes and ‘=1’ indicates that only individuals with diabetes (coded as 1) are included in the study. Note, this is not specified on the CMs to simplify figures. Cachexia is represented by ‘U’, and ‘COPD’ is chronic obstructive pulmonary disease. With respect to the longitudinal DAGs, ‘H’ (history) denotes status before the study, ‘0’ denotes baseline, and ‘1’ represents the first time point after baseline. There may be subsequent time points following this (denoted by the ‘…’), that would require a new set of variables. In the CMs, mortality rates are denoted by dotted lines which point to empty space. Rates with no labels (including mortality rates) may all be distinct. For all rate naming conventions, refer to Appendix model Eqs. (2) (for the Preston et al. and age-structured models) and (3) (for the reverse causation and combined models).

There have been numerous potential explanations proposed for the obesity paradox Examples include reverse causation [9], confounding, selection bias [9, 12], or inaccuracy of BMI to represent body composition [13]. In general, underlying causal mechanisms that have not properly been adjusted for in the analysis may cause bias. Other causal explanations include the fact that obese individuals may receive better medical treatment [14], or are chronic disease specific e.g. obese individuals may be protected from plaque formation on their arteries through a greater mobilization of endothelial progenitor cells [15].

For the purposes of this study, we will define the obesity paradox based on the qualitative results of the Preston et al. study, i.e., the obesity paradox occurs when obese *never-smoking* diabetics have *higher* rates of mortality than normal weight never-smoking diabetics and obese ever-smoking diabetics have *lower* rates of mortality than normal weight ever-smoking diabetics. We also assumed that comparable individuals who are obese or ever-smokers always have higher mortality rates than their normal weight or never-smoking counterparts, respectively. In other words, we only considered biases in study designs (specifically reverse causation or selection bias) as potential explanations, rather than examining situations where we model obesity as actually being biologically protective.

To obtain an unbiased effect measure of BMI on mortality, we can refer to the structure of the DAG from Preston et al. (Figure 2(a)). Overall, if we assume that there are no other sources of bias in the study, and no other common causes of the variables on the DAG, an unbiased effect estimate would require that we adjust for smoking status. Diabetes is a collider or common effect of smoking and BMI, and a mediator in the path between BMI and mortality. Conditioning on a collider creates a spurious association between its causes [16] however, adjusting for smoking removes the bias. To account for the fact that we are conditioning on a mediator (diabetes is a mediator on the path between BMI and mortality), we can assume that there are no additional unmeasured confounders and only consider the controlled direct effect of BMI on mortality i.e., when diabetes is held constant [17]. See Appendix Section A1.3 for a more details on determining what to adjust for in the statistical analysis.

### 4.2 Workflow Summary

We propose the following workflow to simulate epidemiological studies and conduct statistical analyses on CMs derived from DAGs:

1. **DAG and Study Design**. Design or use an existing DAG representing the causal processes related to a given exposure and outcome, and then design an epidemiological study (alternatively, this method can be used to conduct sensitivity analyses on an existing study in which case one would use the existing study design). Using the DAG, determine which variables will be controlled for in the statistical analysis (see Step 5 below). In our analysis, we started with a published DAG [9].
2. **DAG→CM Mapping**. Derive a CM from the DAG using the mapping described by Ackley et al. [1].
  a. Because multiple CMs may correspond to the given DAG, decide the appropriate CM based on the chosen study design and realistic mechanisms for the process of interest. In our CM, we used ODEs with exponential transition rates (i.e., exponential waiting time models). In our models, individuals can transition from never-smoking to ever-smoking, but not back to never-smoking. In general, the research question and hypotheses will guide how to correctly derive a CM from a given DAG since the correspondence between DAGs and CMs is not one to one. [1].
  b. Potentially reduce the state-space for the chosen CM based on the study population and biological processes included (e.g., mortality). In our analysis, the study design conditions on diabetes, so we only track individuals with diabetes, and can therefore simplify the model state-space to only include the diabetic states (as opposed to having a corresponding non-diabetic disease state for each diabetic disease state). Similarly, one could reduce the state-space by not including compartments for individuals who have died (corresponding to the mortality variable in the DAG), but rather only including the unique mortality out-flow rates from each compartment.
3. **Simulation and Sampling**. Simulate the chosen study population using the CM based on predefined ranges of parameter values and initial conditions. In our analyses, we simulated a yearlong longitudinal cohort study among diabetics aged 40-74 (this matched the ages of the population in the observational study) for each sampled parameter set.
  a. Parameter and initial condition values and ranges can be determined based on the mechanism of interest, existing data, the literature, or simply broad ranges that encompass the plausible space of values (as were used in our analysis). Values may be (for example) uniformly sampled from these distributions using Latin Hypercube Sampling (LHS) [18].
  b. Simulation of the study using the chosen CM can be implemented in a variety of ways, e.g. as ODEs (for sufficiently large populations) or as a stochastic model. Among smaller populations, it is often necessary to use a stochastic model, because the stochastic variability not captured by ODEs may lead to unexpected findings.
4. **Generate Simulated Data**. Generate a simulated dataset based on the outcome of interest and measurement details of the study (e.g. number of follow-up time points, variables measured, potential sampling or measurement error that might be of concern). In our case, because individuals were followed up once at the end of the study, we made a single simulated dataset for the entire study (consisting of person-time and the outcome, mortality, by disease state over the course of the year). For simplicity and because we simulated a very large study (1,000,000 individuals), we did not examine issues of sample size or measurement error. After simulating a study, we subsequently calculated person-time (to estimate time at risk for the study population over the course of the study) and incident mortality by disease state.
5. **Analysis and Evaluation**. Run statistical analyses and/or calculate the causal parameter of interest using the simulated data in Step 4. Analyses may include calculation of a single effect estimate and/or a wide range of statistical regression methods (depending on what analyses are of interest/planned for the study). Next, evaluate the results to examine how the causal relationships and parameters included in the model affect potential biases and patterns of interest in the data. In our analysis, for our outcome we calculated mortality rate ratios (MRRs) by dividing normal weight mortality rates by obese mortality rates among individuals within different smoking strata and then assessed whether each given model and study design could recreate the obesity paradox. For an example calculation of MRRs, see Appendix Section A1.17.
6. **Revision and Exploration**. Based on the results of Step 5, potentially alter the study design and/or DAG to explore alternative biases and causal mechanisms, then re-run the workflow. We did this by simulating epidemiological studies assuming different unadjusted study design biases (i.e., reverse causation and selection bias).

In the remainder of this paper, we assess how different underlying causal mechanisms might lead to the obesity paradox to illustrate the utility of this workflow. Example code that we used for our analyses which demonstrates this workflow is available on GitHub: https://github.com/epimath/cm-dag.

### 4.3 Simulating a Longitudinal Cohort Study

We simulated a yearlong cohort study to examine the relationship between obesity and mortality among diabetics aged 40-74. We followed up participants once at the end of the study to calculate person-time and incident mortality by disease state. We started with a population of 1,000,000 people and (for the age-structured models mentioned below) weighted according to their age group distribution in the 2010 United States (US) census [19]. See Appendix Section A1.8 for details on age-weighting for our study population.

### 4.4 Alternative CMs

We used 4 different ODE models to explore how our simulated datasets change with different proposed underlying causal mechanisms. See Figure 2 for all DAGs and corresponding CMs. We began with <u>Model 1</u>, a direct conversion of the published DAG from Preston et al. [9] to a CM. See Appendix Section A1.4 for details on how we converted this DAG and reduced the number of compartments on the CM. After following the workflow for Model 1, we explored other possible mechanisms that might lead to the obesity paradox. The other mechanisms used in this case study were inspired by the Preston et al. study and literature, to evaluate whether they can provide a plausible explanation for the obesity paradox—we note this simulation study cannot provide an actual explanation to the obesity paradox, only test whether particular hypothesized mechanisms can potentially generate the obesity paradox. <u>Model 2</u> incorporated age-varying rates and was age-weighted according to the US census [19]. We split our population into a younger age-group (ages 40-59) and an older age-group (ages 60-74) and simulated the same model within strata of age. See Appendix Section A1.7 for details on how we incorporated age into the DAG and CM. <u>Model 3</u> represents reverse causation due to chronic obstructive pulmonary disease (COPD), a co-morbidity associated with diabetes for which smoking is a risk factor that can induce cachexia (loss of weight and muscle mass) and cause higher mortality rates [20-23] (thereby increasing mortality among a subset of normal weight ever-smokers). Individuals with comorbid diabetes and COPD can transition into an ‘unhealthy’ compartment, *U*. Individuals in *U* have lost weight due to cachexia and also have higher mortality rates than their normal weight ‘healthy’ counterparts (i.e. normal weight ever-smoking individuals with COPD who have not undergone cachexia). See Appendix Section A1.9 for details on the underlying mechanism and how we incorporated reverse causation into the DAG and CM. We note that in this case when mortality is the outcome, reverse causation can more accurately be described as confounding by disease (i.e., disease affects both weight loss and mortality) [24]. However, this type of confounding is often termed ‘reverse causation’ [25, 26] (as it is in Preston et al. [9]), and thus to be consistent with Preston et al. we will refer to it as reverse causation. Finally, Model 4 is a combination of Models 2 and 3. See Appendix Section A1.11 for details on how we incorporated age and reverse causation into the DAG and CM.

In all CMs, once individuals die, they cannot move between disease states and we no longer track them, therefore for simplicity, mortality is treated as an outgoing flow from each compartment and was not included in the set of disease states. Similarly, in the CM diagrams, death rates (mortality) are typically drawn as arrows pointing to empty space, a convention we have also used in this study. In Figure 2, transition rates that are not labeled are assumed to be distinct. For instance, the mortality rates (denoted by dotted lines), are different for each disease state and represent different causal effects on mortality (e.g., obesity, smoking history).

### 4.5 Parameterization of the CM

We aimed to make minimal assumptions about parameter values, to derive generalizable insight into potential mechanisms underlying the obesity paradox. We conducted a sweep of parameters (transition and mortality rates) and initial states (denoted ‘parameter sets’) using LHS [18] to uniformly sample values from predefined ranges [18, 27]. Specifically, we allowed all compartment transition rates to vary from 1% to 20% per year. For example, this results in between 1% to 20% of obese ever-smokers becoming normal weight over the course of the 1 year study. Although 20% is unrealistically high (especially in the general population), we intentionally set a large range of parameter values to ensure that we capture realistic ranges and to see if any extreme scenarios might lead to the obesity paradox. Furthermore, we placed no restrictions on the number of individuals starting in each state and only ensured that the total number of individuals across all disease states equaled the study population at the start of the simulation. See Appendix A1.12 for more details on the calculation of (and alternative methods to derive the) initial conditions. The initial conditions were determined by random sampling from the space of possible proportions of the total population in each compartment (i.e. we randomly chose the initial compartment values conditioned on the total being the correct total population size). We imposed biologically realistic restrictions on the mortality rates such that ever-smokers have a higher mortality rate than their never-smoking counterparts (i.e., within weight strata), and obese individuals have a higher mortality rate than their normal weight counterparts (i.e., within smoking strata). In the age-structured models, older age group mortality rates for a given disease state were determined by multiplying the younger age group mortality rate of the same state by a multiplier between 1 and 2. Finally, in the reverse causation models, we derived the mortality rate in the *U* compartment by multiplying the mortality rate of normal weight healthy ever-smokers with COPD by a cachexia multiplier between 1 and 2 (similar to the age multiplier in Model 2). For instance, the mortality rate for smoking in the older age-group is the baseline mortality rate plus the smoking mortality rate add-on value. This sum is then multiplied by the age-varying mortality multiplier. Note, we incorporate both add-ons and multipliers in our mortality rates to illustrate different simple options for defining rates relative to each other while making minimal assumptions. While these simplifying assumptions ensure that risk factors such as obesity always increase mortality, effects on mortality can be modeled generically as arbitrary different values when there is a causal effect between two states. Overall, each model represents different underlying causal mechanisms and running a model on a given parameter set represents a single simulated study. See Appendix Section A1.13 for more details on sampling transition and mortality rates for each model and Table 1 for all LHS ranges.

**Table 1:** Parameters and Latin Hypercube Sampling Ranges for All Models

| Parameters | Range | Models |
| --- | --- | --- |
| Normal weight never-smoking to obese never-smoking | 0.01 to 0.2 | All models |
| Obese never-smoking to normal weight never-smoking | 0.01 to 0.2 | All models |
| Smoking initiation rate | 0.01 to 0.2 | All models |
| Normal weight ever-smoking to obese ever-smoking | 0.01 to 0.2 | All models |
| Obese ever-smoking to normal weight ever-smoking | 0.01 to 0.2 | All models |
| COPD incidence rate | 0.01 to 0.2 | Model 3 and combined model |
| Normal weight ever-smoking with COPD to obese ever-smoking with COPD | 0.01 to 0.2 | Model 3 and combined model |
| Obese ever-smoking with COPD to normal weight ever-smoking with COPD | 0.01 to 0.2 | Model 3 and combined model |
| Cachexia initiation rate | 0.01 to 0.2 | Model 3 and combined model |
| Baseline mortality rate | 0.01 to 0.1 | All models |
| Add-on for smoking | 0 to 0.1 | All models |
| Add-on for obesity | 0 to 0.1 | All models |
| Age-varying mortality multiplier | 1 to 2 | Model 2 and combined model |
| Add on for COPD | 0 to 0.1 | Model 3 and combined model |
| Cachexia ($U$) multiplier | 1 to 2 | Model 3 and combined model |

#### 4.5.1 Data Generation and Statistical Analysis

After running each model with 10,000 randomly sampled parameter sets [28], we calculated person-time and incident deaths per compartment for each study (i.e. for each model and sampled parameter set). See Appendix Sections A1.14 and A1.16 for more information on these calculations. Next, we calculated MRRs comparing normal weight to obese individuals within smoking strata to measure the effect of BMI on mortality. As mentioned, to recreate the obesity paradox (as per [9]), the MRRs from a simulated dataset (i.e., study) must simultaneously show normal weight never-smokers with *lower* mortality rates than their obese counterparts, and normal weight ever-smokers with *higher* mortality rate than their obese counterparts.

In Model 1, we measured all compartments and calculated the MRRs directly from the simulated data. In Model 2 (age), we initially did not adjust for age as a confounder. Rather, MRRs were calculated by taking the sum of incident deaths divided by the sum of person-time for a given disease state across age-groups. As a sensitivity analysis, we adjusted for age by externally standardizing the MRRs to the reference (obese) group [29]. Finally, in Model 3 (reverse causation), our study design did not initially adjust COPD or related complications (i.e., cachexia). Therefore individuals with COPD were measured together with ever-smokers (e.g., in our study population, all normal weight individuals with COPD including those with cachexia were measured together with normal weight ever-smokers). The MRRs were calculated in the same way as we did for Models 1 and 2. Therefore, we initially did not adjust for COPD or cachexia. As a sensitivity analyses, we adjusted for reverse causation by excluding all individuals with COPD (including those with cachexia) at baseline (and then ran the study for 1 and 5 years). Finally, in the combined model, we ignored age, COPD, and cachexia in our initial analysis, and then adjusted for age only, COPD only and finally, age and COPD.

All simulations and analyses were conducted in R version 3.3.3 [30]. Compartmental models were run using the ode function from the ‘deSolve’ package which uses lsoda (an adaptive stiff/nonstiff integrator) [31].

## 5 Results

Overall, we found that not adjusting for study design bias in our CMs resulted in the obesity paradox. See Table 2 and Figure 3 for all results.

**Table 2:** Results from All Analyses

| Models | Unadjusted Analysis | Adjusted Analysis |
| --- | --- | --- |
| Model 1: Published DAG | **No obesity paradox** | NA |
| Model 2: Adding in age-varying mortality rates | **Obesity paradox occurs** – when there are more younger obese or more older normal weight individuals (selective survival bias) | Adjusting for age stops the obesity paradox from occurring |
| Model 3: Reverse causation | **Obesity paradox occurs** – more than in Model 2 because the mechanism directly affects normal weight individuals (reverse causation bias) | Excluding those with COPD at baseline stops the obesity paradox from occurring (for the 1 year, but not the 5 year study) |
| Model 4: combined | **Obesity paradox occurs** – Predominant mechanism is reverse causation | Interactive effects between biases |

**Figure 3:**
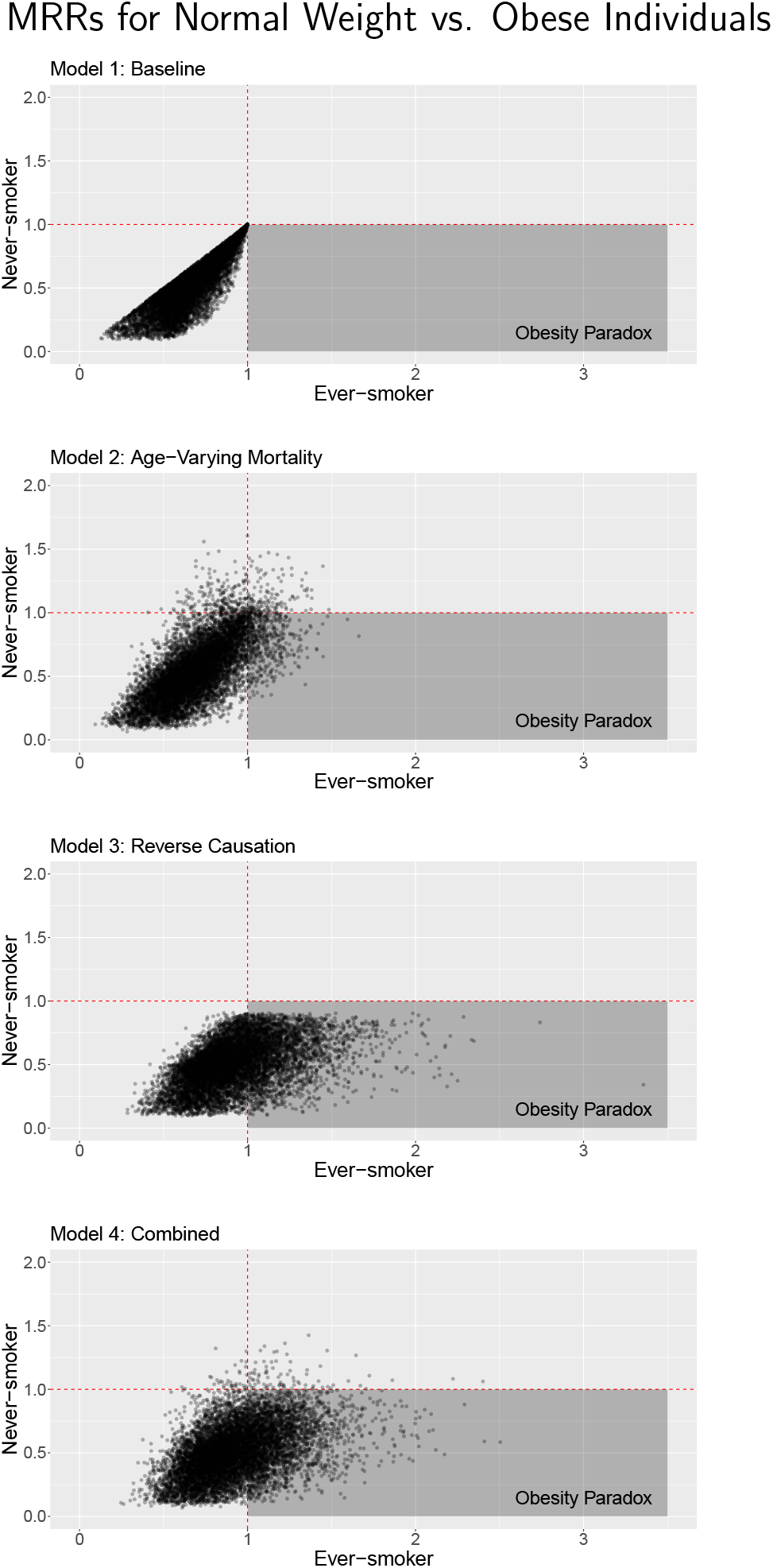
Results from all model runs and parameter sets. Each plot represents the MRR (mortality rate ratio) comparing normal weight individuals to obese individuals for never-smokers against the corresponding MRR for ever-smokers for each of the 10,000 LH-sampled parameter sets with each point representing a single simulated study. The obesity paradox occurs when obese never-smoking diabetics have *higher* rates of mortality than normal weight never-smoking diabetics and obese ever-smoking diabetics have *lower* rates of mortality than normal weight ever-smoking diabetics. By row: 1. Preston et al. [9] 2. adding in age-varying mortality rates; 3. reverse causation and 4. combined model.

In Model 1, we did not see the obesity paradox because the MRRs from the simulated data were simply the ratio of the CM mortality rate parameters (see Figure 3(a)). For instance, the ever-smoker MRR is just the mortality rate of normal weight diabetic ever-smokers (*NWDS*) divided by the mortality rate of obese diabetic ever-smokers (*ODS*) (see Appendix Section A1.17 for more details). Due to the structure of Model 1 and the restrictions we placed on the parameter values, mortality rates for normal weight individuals were always lower than (or at the very least equal to) their obese counterparts therefore, all ever-smoking MRRs were ≤1. Overall, Model 1 cannot simulate a protective effect of obesity on mortality among diabetic ever-smokers.

Next, in Model 2, the obesity paradox did occur in a subset of studies (See Figure 3(b)). Overall, among model runs that resulted in the obesity paradox, there were generally either more younger individuals in the obese ever-smoking compartment and/or more older individuals in the normal weight ever-smoking compartment. This caused the mortality effects of age to counterbalance those of obesity, resulting in the obesity paradox. In other words, for the obesity paradox to occur, age-varying mortality must be sufficiently high and work together with the relative age distribution of individuals across disease states. This is analogous to selective survival bias in which obese ever-smoking individuals are more likely to die before they reach older ages, thus there would tend to be more older normal weight ever-smokers than older obese ever-smokers. An illustration of this is the trade-off between the proportion of old vs. young individuals who are in the obese diabetic ever-smoking (*ODS*) compartment and the relative mortality rate of normal-weight diabetic ever-smokers (*NWDS*) vs. *ODS* (shown in Figure 4). The majority of parameter sets that resulted in the obesity paradox show the proportion of individuals in the older age-group among all *ODS* is < 50%. Additionally, the effect of obesity on mortality is relatively low (i.e., the *NWDS* mortality rate is consistently similar to the *ODS* mortality rate in the parameter sets that resulted in the obesity paradox). Finally, as the proportion of individuals in the older age group increases, the effect of obesity on mortality decreases even more in model runs that resulted in the obesity paradox. This is analogous to obesity becoming less risky as individuals age [32]. In the age-standardized sensitivity analysis, no runs resulted in the obesity paradox (results not shown; similar to the baseline model).

**Figure 4:**
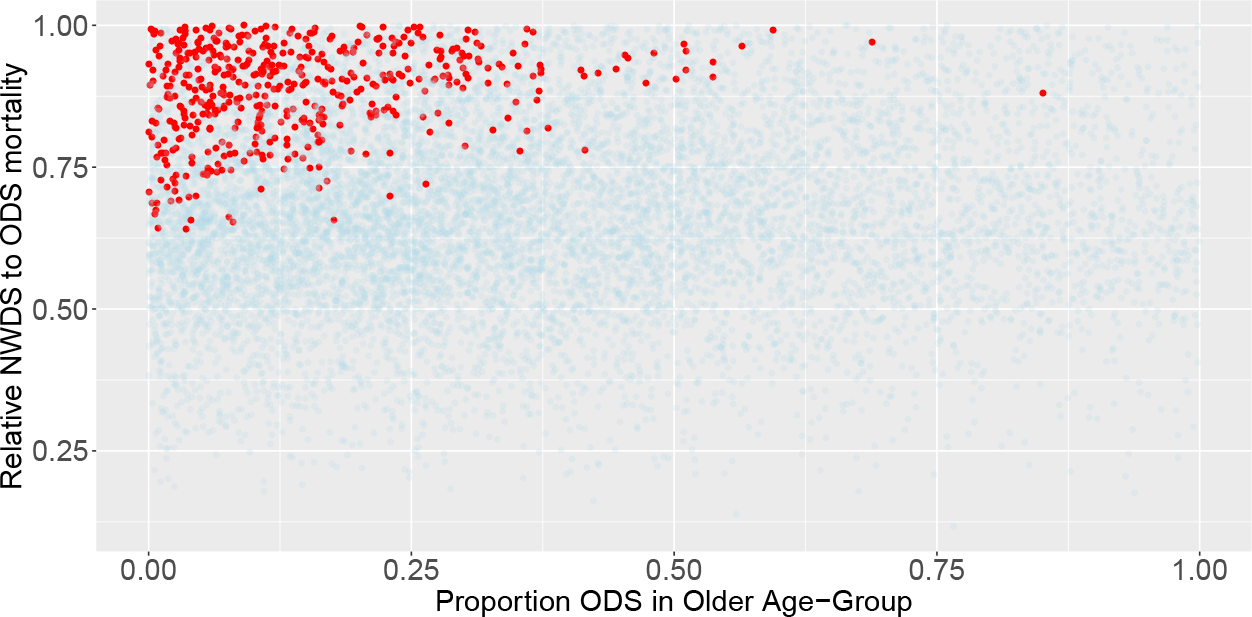
Age-weighting and relative mortality among Model 2 runs. The proportion of obese diabetic ever-smokers (*ODS*) who are old at the beginning of the simulation is displayed on the x-axis e.g., if equal to 0.5, half of the individuals in ODS are in the older age group and half are in the younger age group. The relative mortality of normal diabetic ever-smokers (*NWDS*) to ODS is displayed on the y-axis e.g., if equal to 0.5, the *NWDS* mortality rate would be half of the ODS mortality rate. Parameter sets that resulted in the obesity paradox are in red and sets that did not result in the obesity are in blue. When *NWDS* mortality is close to ODS mortality, having ODS be primarily younger can counterbalance the higher mortality due to obesity.

In Model 3 (compared with Model 2), more runs resulted in the obesity paradox (Figure 3(c)). This is due to the fact that the reverse causation mechanism differentially affects normal weight ever-smoking individuals (compared with obese ever-smoking and normal weight never-smoking individuals). Therefore, the obesity paradox depends on (1) the relative obese and normal weight mortality rates (for both healthy and unhealthy individuals) and (2) the distribution of individuals in healthy and unhealthy compartments. On the other hand, in Model 2, age-related mortality affects normal weight and obese individuals as well as ever-smoking and never-smoking individuals in the same manner and thus relies on the population distribution across more compartments i.e., both age groups in ever-smoking obese and normal weight and never-smoking obese and normal weight compartments. Because healthy and unhealthy normal weight ever-smokers are measured together in our observational study, the unhealthy mortality rate increases the combined (healthy and unhealthy) normal weight ever-smoking mortality rate such that the overall normal weight ever-smoking mortality rate is higher than the obese ever-smoking mortality rate and the obesity paradox occurs. When examining results among ever-smokers only, the weighting of the overall normal weight mortality rate is revealed in the relative proportion of individuals starting in different disease states (Figure 5). For instance, for runs in which the obesity paradox occurs, the relative mortality rate of individuals who are unhealthy compared to those who are obese ever-smokers increases when fewer normal weight individuals start in the unhealthy compartment.

**Figure 5:**
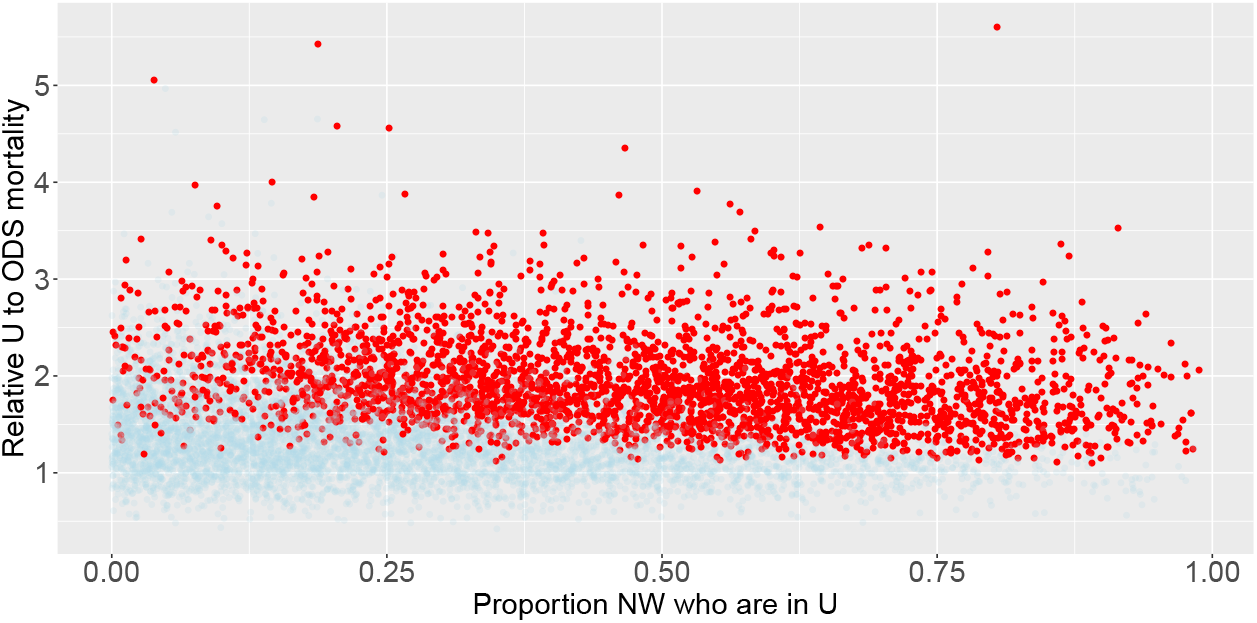
Age-weighting and relative mortality among Model 3 runs. The proportion of normal weight individuals who are unhealthy at the beginning of the simulation is displayed on the x-axis. The relative mortality of unhealthy individuals to obese diabetic ever-smokers (*ODS*) is displayed on the y-axis i.e., if equal to 2, the *U* mortality rate would be twice of the *ODS* mortality rate. Parameter sets that resulted in the obesity paradox are in red and sets that did not result in the obesity are in blue. As *U* mortality becomes substantially higher than *ODS* mortality, having fewer individuals starting in *U* is sufficient to counterbalance the higher mortality due to obesity.

The results from our sensitivity analyses reveal that excluding individuals with COPD at baseline reduced the number of model runs that result in the obesity paradox to 1 (compared with 3,114 in the unadjusted version). If we run the study for 5 years to evaluate whether the obesity paradox might emerge over time, only 82 model runs resulted in the obesity paradox (results not shown). This highlights the importance of inclusion and exclusion criteria in an initial study population in recreating the obesity paradox.

Finally, in the combined model, we found that reverse causation leads to the obesity paradox substantially more than age-weighting (selective survival). This is evidenced by the fact that when we adjust for age only, the obesity paradox still occurs in 97.1% of the runs in which it originally occurred (3017/3107), while if we adjust for reverse causation only, the obesity paradox occurs in 12.1% (376/3107) of the runs in which it originally occurred. The results from our sensitivity analyses reveal that when we control for both age and COPD, the obesity paradox is avoided almost completely. Interestingly, when we standardize age or exclude individuals with COPD only, certain parameter sets that did not previously result in the obesity paradox, now demonstrate the obesity paradox. This indicates a ‘two wrongs make a right’ interactive effect between these two biases: for instance, if the majority of normal weight individuals are younger, this might counteract the effects of a high proportion of individuals starting in *U* in the unadjusted model, but if we adjust for age only, the high proportion of individuals starting in *U* may result in the obesity paradox.

## 6 Conclusion

We have developed a workflow that can be used to explicitly examine the underlying conditional independencies of DAGs. This method provides a systematic way to quantitatively simulate and evaluate bias and provide insight into the causal relationships between variables in a study. Our workflow can be applied to nearly any study question assuming standard assumptions (e.g., no faithfulness violations on DAGs [33]). Previous work by Ackley et al. [1], provided a basic mapping from DAGs to CMs with simple examples (e.g., confounding, collider bias). We extended this work by noting opportunities to reduce the state-space (e.g., by removing diabetic compartments) and creating a workflow to use this mapping to simulate epidemiological studies and assess potential biases affecting study results. Finally, we applied this workflow by using and adapting a published DAG [9]. Importantly, although we made explicit parametric assumptions (stronger than those used on a DAG), we allowed for a large range of transition rates and initial conditions such that we only made minimal quantitative assumptions (e.g., we assumed that the maximum number of individuals that transition between a given disease state to another is 20% over the course of the year). In future work, parameter values could be informed using study– and setting–specific data if available.

Modeling DAGs and conducting simulated studies can provide insight into how to design sound observational studies and analysis plans. For instance, if results from a simulated study don’t match expected results, this may imply the existence of unmeasured and/or unadjusted covariates, or interacting biases as we found in our obesity paradox simulation study. Although in this case, traditional analyses using DAGs would have likely found the same main sources of bias (i.e., unadjusted covariates), our method also identified some additional biases that would not have been easily identified using traditional methods (e.g., interaction between age and reverse causation, or excluding individuals with COPD at baseline and then running the study for 5 years shows that biases that were initially adjusted for could re-emerge over time depending on the length of follow up). Overall, we simulated epidemiological study data in a structured manner based on the conditional independencies of DAGs to test different hypotheses. Additionally, this framework allows us to explore and simulate these biases interactively and over time, observing (for example) how different measurement times, sampling designs, etc. might affect potential biases or impact which variables are most critical to measure.

We successfully recreated the obesity paradox by deriving a compartmental model from a published DAG [9] and then incorporating two different unadjusted biases. In Model 1, we found that direct conversion of the published DAG was not able to recreate the obesity paradox. In Model 2, we incorporated age-varying mortality and found that the relative proportion of individuals in different age groups across disease states can create a selective survival bias causing the obesity paradox. In Model 3, we found that reverse causation caused by an unmeasured disease state can more effectively cause the obesity paradox compared with the age-varying mortality model. The reverse causation mechanism was more effective because it differentially affected normal weight ever-smoking individuals (compared with obese ever-smoking and normal weight never-smoking individuals). Finally in the combined model, we observed how different biases can interact to cause or prevent the obesity paradox from occurring. Overall, adjusting for biases in these models (sensitivity analyses) made the obesity paradox nearly non-existent, indicating that incorporating bias and not adjusting for it correctly is required to recreate the obesity paradox (assuming the protective effect of obesity is not truly present, and that we have sufficient sample size). Ultimately, even with very general parameter assumptions for our model, we were able to derive insight into what causal mechanisms may drive the obesity paradox. In the literature, both selection bias [34] and reverse causation [9, 25] have been suggested as potential causes of the obesity paradox in previous studies. Our results suggest that, over the parameter ranges we explored (and for this specific study design and simulated population), reverse causation may be more effective than age-related mortality at generating the obesity paradox, however more realistic, specified modeling studies are needed to further explore these issues. Moreover, other study biases likely co-exist and depend on the specific dataset and study design. However in general, in situations where only a limited number of variables can be measured e.g., due to logistical constraints, our workflow could be used to identify which biases are more important to account for (i.e., reverse causation in our analyses) and therefore which variables to measure.

Overall, our analyses were primarily meant to illustrate the utility of our workflow and further study would be needed (e.g. with more potential causes or biases, and estimation or sampling of parameters from data) to thoroughly investigate the causes of the obesity paradox. Therefore, we drew our inspiration from the Preston et al. study [9], but did not aim to recreate their results directly. Depending on the goals of the study, our workflow can be used to quantitatively recreate the results of existing studies which can be used to more precisely derive new insight into which study design biases are predominant, how biases might interact, or what combinations of factors lead to a specific scientific conclusion.

Although DAGs alone can (and are often used to) indicate the presence or absence of bias, they don’t include the magnitude or direction of effects (although one may be able to infer direction based on the type of bias). There have been a number of recent methods and software developed to allow the incorporation of different parametric assumptions and direct simulation of DAGs [35–37]. While these methods will in many cases likely lead to the same results as our workflow (and in some cases may be equivalent to a type of CM), there are a few key advantages of translating DAGs into CMs. First, CMs can include transitions between disease states that aren’t the encoded on the DAG, by setting transition rates equal to each other for different compartments (see Figure 1 for an example). Additionally, DAGs are restricted by the limitations of statistical analyses e.g. no interference between units assumption [38], while CMs can be expanded to examine processes that may violate this assumption, such as infectious disease processes, as was illustrated in [1]. There are also limited options to encode different functional form relationships between variables in DAGs. For instance, DAGs do not have a formal way to represent effect modification [39]. Because CMs are more explicit in encoding the natural history of individuals in a dynamic way, they represent causality differently from DAGs. For a given mechanism under consideration, CMs (1) define the steps sufficient to be able to progress to a given outcome e.g., cachexia, and (2) track the dynamics of the number of individuals in disease states over time. Therefore, CMs represent a population akin to what would be seen in a study or in the real-world, which can be advantageous but also requires more detailed information or assumptions to construct. Thus, using CMs can help to develop understanding and intuition for the process under consideration. This allows the CM to act as a sort of ‘population laboratory’ in which different causal processes and mechanisms can be explored and simulated. Overall, our workflow can be used to simulate epidemiological studies while allowing for the simultaneous incorporation of different types of bias and different underlying causal mechanisms. Although this may also be possible through the direct simulation of DAGs, CMs are highly flexible and intuitively causal. A useful direction for future work will be to compare these different DAG simulation approaches and evaluate the different aspects of a given study/dataset each can be used to explore.

Notably, the sufficient-component causes framework has been incorporated into DAGs and can provide detail about underlying mechanisms by illustrating how causes interact for a given outcome to occur [40]. This is akin to detailing the individual steps sufficient for a given outcome in CMs, though this method does not require explicitly separating all steps in the sequence of events for a given outcome. One advantage of this method is that it makes relationships between causes more explicit e.g., conditionally independent or synergistic. In a CM, these relationships may only appear implicitly in the transition rates. Additionally, this method requires the inclusion of all possible sufficient causes, while a CM only includes the mechanisms under consideration.

Limitations of this study include the fact that the DAGs we use are overly simplified (despite our use of a published DAG) and do not represent the complete state of knowledge about the relationships between variables relevant to the study question. We decided to use relatively simple DAGs to more effectively illustrate our workflow. It is simple to make more realistic DAGs by adding additional demographic characteristics e.g. race, socioeconomic status, access to medical treatment and including these would simply require vectorizing our equations further (as we did for the extension from Model 1 to Model 2). However, since we are not fitting these models to study data that includes these variables, we would have added more parameters to our models without truly adding any information. Because each new DAG variable doubles the number of equations in the CM, this would add complexity without insight. We aimed to strike a balance between realism and parsimony in our models to isolate and examine the qualitative effects of individual causal mechanisms of interest. For instance, the effects of race may counteract the effects of age leading to overly complicated results (i.e. identifiability issues may obscure the larger point). A potential future direction is to construct larger DAGs from the literature and make simplifying assumptions to reduce the corresponding CM’s dimensionality (such as including only one variable among a collinear set). For instance, suppose both BMI at baseline and BMI history are included on a given DAG, one could assume that history is a proxy for baseline BMI among e.g. adults [41] and collapse these two variables into a single BMI variable. The robustness of results to this simplifying assumption can also be explored using our workflow. Relatedly, our workflow could also be used to identify which variable(s) on a DAG are sufficient or necessary to replicate a particular pattern in the data (e.g., by systematically removing variables and simulating the results). Finally, individually-based models may be used for study questions requiring more detailed demography. Furthermore, individually-based models can be used to explicitly track individuals and their transitions between disease states. Another weakness is that our crude estimate of person-time (see Appendix Section A1.14) will not work if the dynamics of the model are very fast. It is possible to calculate person-time precisely by tracking the flows in and out of compartments separately. Lastly, the exponential transitions between disease states used in our CMs were chosen for the sake of simplicity and may not be realistic. To address this simplifying assumption, we also tested incorporating a gamma distributed transition rate from normal weight individuals with COPD to cachexia in the reverse causation model (see Appendix Figure A3 for model schematic). Ultimately, we found that incorporation of this different functional form reduced the number parameter sets that resulted in the obesity paradox in the 5-year adjusted analysis (i.e., after excluding individuals with COPD at baseline and running the study for 5 years), however the overall qualitative conclusions did not change. More generally, changing the formulation of our parameters (e.g., by making the parameters time varying or alternatively, gamma distributed) is an important topic for exploration, although it would be less likely to affect the overall qualitative conclusions of our particular simulation study, because the factors leading to the obesity paradox (i.e., the restrictions we placed on the relative values of mortality rates and how we measured the variables in our study) would still be the same. Broader distribution types beyond gamma distributions are also possible to incorporate into ODE models (e.g. Erlang mixture distributions and more general families of dwell time distributions [42]), or a more generalized stochastic framework can be used.

Strengths of this study include the methodological contributions to using CMs in conjunction with DAGs to understand patterns seen in the data. We extended the mapping that Ackley et al. developed [1] and proposed a method for comparing simulated data with epidemiological study data. This method can be expanded for different types of epidemiological analyses and can also be used for different purposes e.g. relaxing statistical assumptions, multifaceted sensitivity analyses or exploring counterfactual scenarios. We were able to show that the initial, simple DAG presented in the Preston et al. paper did not on its own reproduce the obesity paradox and then proposed alternative mechanisms and DAGs that could recreate the obesity paradox. Furthermore, we gained insight into what hypothetical causal mechanisms could result in the obesity paradox with limited data informing our model. Additionally, conducting the random sweep (i.e. LHS) of the parameters and initial conditions allowed us to account for uncertainty and draw general qualitative conclusions about the structure of the model and its effects on our statistical results. Ultimately, our workflow can help explicate causal mechanisms to explore whether or not DAGs are valid representations of hypotheses in question even when data is limited. Additionally, CMs derived from DAGs can be used as a testing ground for competing causal mechanisms to determine which ones can most closely explain patterns seen in observational study data. This represents a departure from the standard paradigm of fitting CMs to epidemiological data, where instead, here we operationalize causal relationships depicted on the DAG to simulate epidemiological study data.

Additional future research can include other statistical analyses on simulated data. For instance, a Poisson regression model (for count data) can calculate MRRs and can be useful if conditioning on multiple variables (see Appendix A1.17.1). Alternatively, simulated data can be individuated and other types of regression models can be run. Model parameters can be tuned to quantitatively recreate specific datasets which might be useful for gaining insight into specific study results or a specific target population. Additionally, model parameters can be informed directly from data. For instance, see Appendix Section A1.17 for notes on how to parameterize the mortality rates from data. Similarly, the data collection process itself can be simulated in the compartmental model, allowing one to assess how issues such as measurement error or insufficient power might affect the relationships reflected in the DAG. Finally, the DAG-derived CMs could also be linked with real-world data to accomplish the parameter estimation/inference step itself (i.e. without modeling additional statistical analyses).

Overall, we presented here a new utility for CMs derived from DAGs: testing hypotheses to understand patterns seen in study data. We also proposed a method to compare simulated data with epidemiological study data that can be used to test competing hypotheses. We used our method to determine that a DAG from the literature was not complete and could not recreate the obesity paradox by itself. We therefore simulated two alternative causal mechanisms and derived corresponding DAGs that could recreate the qualitative results of the study. Ultimately, simulating study data by operationalizing the causal relationships on DAGs can provide insight into how to design sound observational studies and analysis plans.

## Data Availability

The datasets and code used for the current study are available from the corresponding author on reasonable request. Example code used for the analysis is available on GitHub.

https://github.com/epimath/cm-dag

## 7 Author Contributions

JH and MCE came up with the research plan. JH analyzed the data and wrote the paper. MCE provided guidance throughout the entire process and provided key edits and feedback on the manuscript.

## 8 Acknowledgements

We would like to thank Dr. Jon Zelner Department of Epidemiology University of Michigan, for his helpful comments and advice on the analysis, Dr. Nancy Fleischer Department of Epidemiology University of Michigan, for her helpful advice about DAGs, and Consulting for Statistics, Computing and Analytics Research (CSCAR) University of Michigan, for their useful advice on the analysis. We would also like to thank Drs. Joseph Eisenberg, Rafael Meza, and Veronica Berrocal for their thoughtful comments on the dissertation version of this manuscript.

## 9 Data Accessibility

Example code demonstrating the workflow is available on GitHub: https://github.com/epimath/cm-dag.

## 10 Funding

This work was supported by the National Institute of General Medical Sciences [Grant Number: U01GM110712].

## A1 Appendix

### A1.1 Brief Overview of Compartmental Models

Compartmental models (CMs) typically decompose all of the members or elements of a system (in this case individuals in a population of interest) into distinct compartments based on their current state (e.g. healthy vs. diseased). Each compartment represents a distinct state and all individuals within a given compartment are typically treated homogeneously in the compartmental framework. In the simplest form of compartmental modeling, in- and outflows to each compartment are treated as Poisson processes (with exponential waiting times), commonly modeled deterministically using linear ordinary differential equations (ODEs). Linear compartmental models have proved enormously useful in a range of application areas, particularly in drug pharmacokinetics and toxicology, where they are commonly used in evaluating drug half lives, dosing regimens, etc. [7]. Compartmental modeling (both linear and nonlinear) is also a regularly used tool in public health for examining disease transmission, ecological processes, and disease progression at the population level [3, 4, 7, 43]. For a more in-depth introduction to compartmental models and their uses, the reader is referred to [7, 43].

### A1.2 Compartmental Model and Directed Acyclic Graph Comparison

See Table A1 for a general comparison between Directed acyclic graphs (DAGs) and CMs.

DAGs are non-parameterized causal diagrams used to graphically map causes and effects to aid in designing epidemiological studies. DAGs summarize the complete set of known relationships between variables relevant to a given study question [1, 44, 45]. A necessary precursor for a DAG to be considered causal is that all known and unknown (if assumed to exist) common causes of any pair of variables on the graph must also appear [1]. Once relationships between variables are synthesized, a researcher can identify what must be measured and/or controlled for to eliminate confounding and selection bias [16, 45]. On DAGs, statistical associations between variables may be produced by (1) cause and effect (unbiased), (2) common causes (confounding bias), (3) common effects (collider bias) [16]. DAGs are therefore used to separate associations due to causality versus those due to bias. DAGs can be represented in a range of different forms including structural equation models or ordinary differential equations as in the case of this paper. Figure A1 shows illustrations of how associations are formed on a DAG. The work flow to structure and then determine which variables to adjust for on DAGs is described elsewhere e.g. [39, 44, 46, 47]. Once assumptions about the causal relationships between variables are made explicit and potential confounders and/or colliders are revealed, a study design and statistical analysis plan can be created such that an unbiased effect estimate of a given exposure on outcome can in principle be calculated.

**Table A1:**
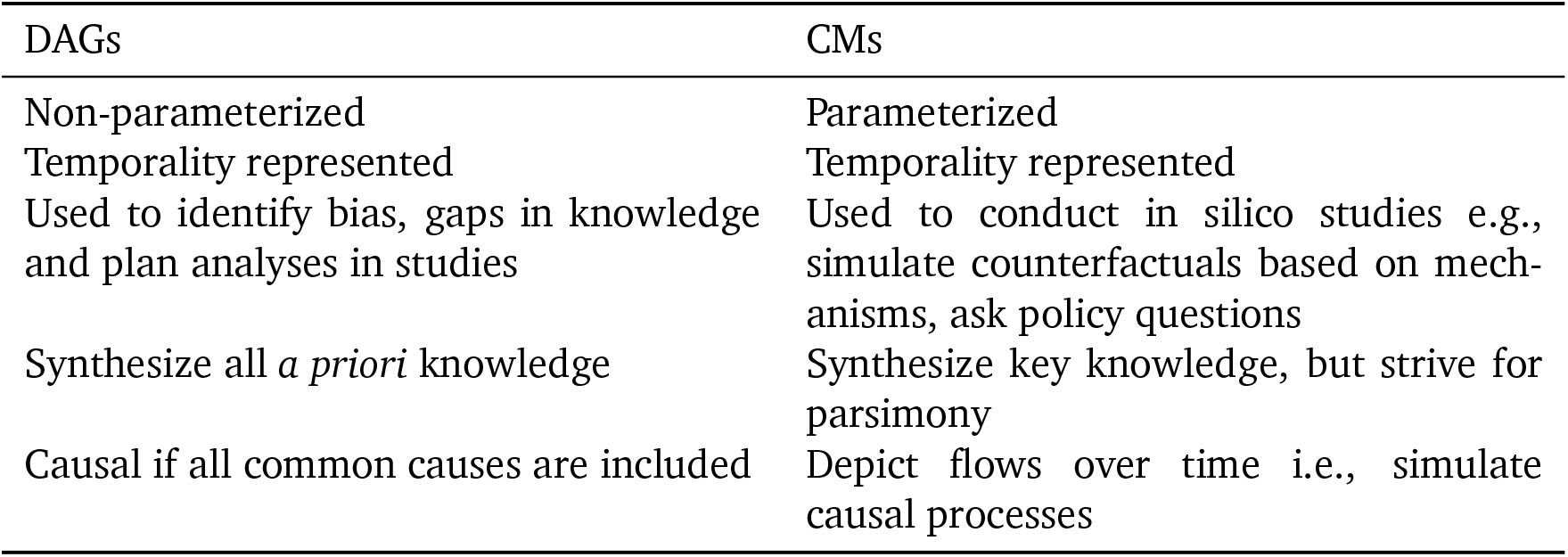
DAGs vs. CMs

**Figure A1:**
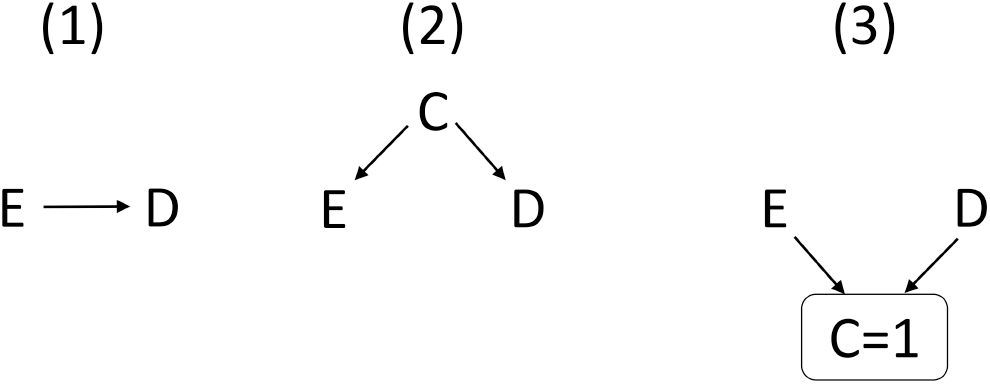
The sources of association between variables become evident on a DAG. *E* is the exposure, *D* is the outcome or disease, and *C* is the covariate (1) cause and effect, (2) common causes or confounding, and (3) common effect or collider bias e.g. selection bias. Here, ‘C=1’ indicates that only individuals with C (coded as 1) are included in the study.

CMs simulate parameterized flows between disease states over time and are themselves a form of causal diagram [1, 2, 48]. Specifically, CMs can be used to explicitly simulate mechanisms underlying disease transmission or disease progression and are often fit to population level data [3, 4]. Unlike causal DAGs which must in principle include all common causes, CMs often must balance realism with parsimony, and often only include the causal processes most relevant to the hypothesis [49].^1^

Once a model schematic is created, it may be converted into ordinary differential equations (ODEs) or simulated stochastically for smaller sample sizes. Data can then be integrated from a variety of sources as model inputs. For instance, data can inform the number of people which start in each disease state (initial conditions) or the transition rate parameter values. Furthermore, model outputs can be fit to a variety of data types [50], such as an epidemic curve or cancer incidence time-series[3, 51]. A fitted model can then be used to (1) estimate transition model parameters and initial conditions, (2) determine which parameters should be measured in future field studies, and (3) examine counterfactual scenarios when data collection is untenable due to ethical constraints or limited resources.

### A1.3 Model 1: Determining What to Adjust for on the Preston et al. DAG

To determine what to adjust for in a statistical analysis, we can refer to the structure of the DAG from the observational study (Figure 2(a)).

Diabetes is a collider or a common effect of smoking status and BMI. The study is conditioned on diabetics (denoted by the box around diabetes on the DAG, and ‘=1’ which indicates that only individuals with diabetes (coded as 1) are included in the study). Conditioning on a collider creates a spurious association between its causes (in this case: smoking status and BMI) also called selection bias [16]. Additionally, diabetes is a mediator on the pathway from BMI to mortality. Conditioning on a mediator typically causes bias when there are unmeasured confounders i.e., between mediator and outcome or exposure and mediator. However, for simplicity, we will assume that there are no additional unmeasured confounders. Even though smoking confounds the association between mediator and outcome (and mediator and exposure), it is measured and we will adjust for it. Other issues related to conditioning on a mediator may arise due to exposure-mediator interaction i.e., if the effect of BMI on mortality is affected by diabetes status. This is addressed by the fact that we will only consider the controlled direct effect of BMI on mortality i.e., when the mediator value is held constant [17]. Next, smoking status is a common cause of BMI and mortality and therefore confounds their association. If we assume that there are no other sources of bias in the study, and no other common causes of the variables on the DAG, an unbiased effect estimate of BMI on mortality would require that we adjust for smoking status. For instance, we will estimate the effect of BMI on mortality in separate smoking strata to remove the spurious associations. We would therefore expect that examining the association between BMI and mortality in a population of diabetics among ever-smokers and then separately among never-smokers would remove the bias and the protective effect of obesity on mortality. However, this was not found to be the case in the Preston et al. study [9]. Therefore, either other biases exist and are not evident due to an inaccurate DAG or incorrectly categorized variables, or obesity truly is protective against mortality among ever-smoking diabetics.

The work flow to structure and then determine which variables to adjust for on DAGs is described elsewhere e.g. [39, 44, 46, 47].

### A1.4 Deriving a Corresponding CM from the Preston et al. DAG

The DAG of interest in this analysis is taken from Preston et al. [9], shown in Figure 2(a). We initially operationalized the causal relationships between variables from the Preston et al. DAG (Figure A2 (left)) by creating a corresponding CM (Figure A2 (right)), using the method of Ackley et al. [1]. Note, we initially did not condition on diabetes (as per Preston et al.) so that we could simulate the entire population, and not just the study population. See Appendix Equations (1) for the ODEs of the full model. We note that these ODE equations represent the deterministic form of Markov exponential waiting time models that track population flows between states. They assume homogeneous populations within each state.

**Figure A2:**
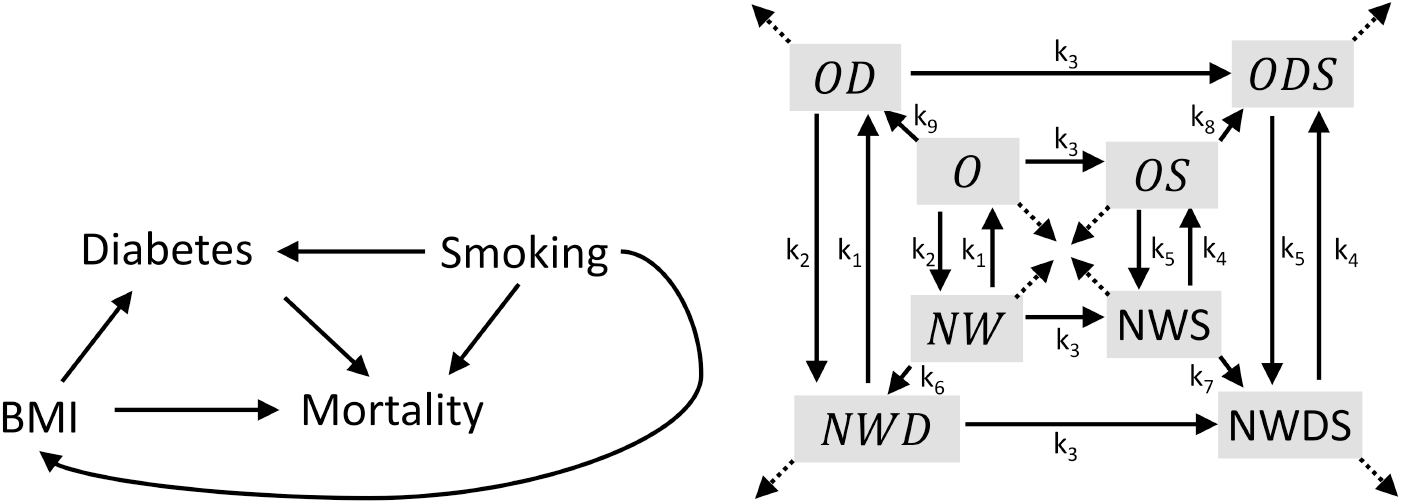
(left) DAG representing the obesity paradox from Preston et al. [9] (right) CM: Schematic of the single age group compartmental model diagram corresponding to the DAG. *NW* represents normal weight individuals; *O* represents obesity; *D* represents diabetes, and *S* represents smoking. Individuals in any given compartment can die. Each arrow represents flows between states and rates that are equal to each other have the parameter. For instance, diabetes status does not affect the rate at which an individual transitions from obese to normal weight, therefore *OD* to *NWD* and *O* to *NW* have the same rate. We specify where transition rates are the same between compartments by labeling the model schematic accordingly and using the same parameter to represent equal rates in the equations. Mortality rates are denoted by dotted lines. Rates with no labels (including mortality rates) may all be distinct. For all rate naming conventions, refer to Appendix Equations (1).

We enumerated disease states based on all possible combinations of random variables appearing in the DAG, i.e. there are 2^4^ possible states, since we have 4 binary variables: diabetes, obesity, smoking, and mortality. Once individuals die, they cannot move between disease states and we no longer track them, therefore to reduce the dimensionality of our model, mortality is visualized as an outgoing flow from each compartment and was not included in the set of disease states. This effectively reduced our model to 2^3^ possible states. However, because we use cumulative mortality for our outcome calculations, we still tracked cumulative incident mortality from each compartment in the code (by multiplying the mortality rate of a given compartment by the number of individuals in that compartment). Next, we included all biologically plausible transitions between states. For instance, an individual can become an ever-smoker, but cannot return to being a never-smoker, also a diabetic individual cannot become non-diabetic.

Since the study population is conditioned on diabetics, we further simplified our model to only include diabetic compartments. This step reduced our model to 2^2^ possible states. See Figure 2(b) for the simplified model schematic and Appendix Equations (2) for the ODEs.

### A1.5 Obesity Paradox Model 1 Full Equations

Below are the ODEs used to simulate the flows between disease states for the full model derived from the Preston et al. DAG. The model schematic is shown in Figure A2 (right). The model equations are given by:

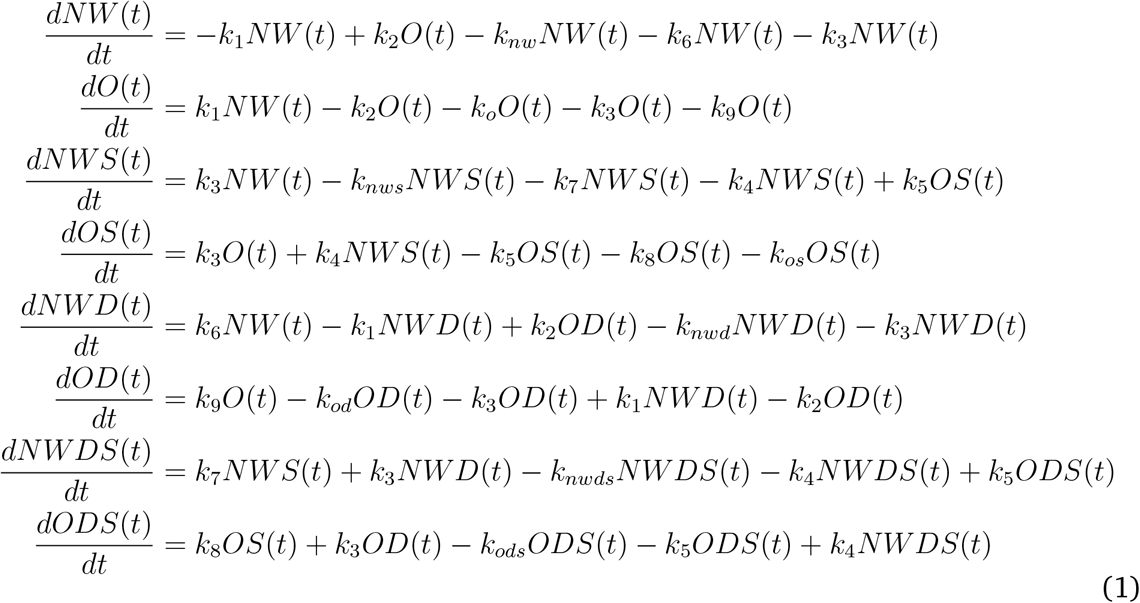

where *NW* are normal weight non-diabetic never-smokers, *O* are obese non-diabetic never-smokers, *NWS* are normal weight non-diabetic ever-smokers, and *OS* are obese non-diabetic ever-smokers. The corresponding compartments for diabetics are *NWD, OD, NWDS*, and *ODS*. *k*_1_ is the rate at which normal weight never-smoking individuals become obese. *k*_2_ is the rate at which obese never-smoking individuals become normal weight. *k*_3_ is the smoking initiation rate. *k*_4_ is the rate at which normal weight ever-smoking individuals become obese. *k*_5_ is the rate at which obese ever-smoking individuals become normal weight. *k*_6_ through *k*_9_ are the diabetes initiation rates for normal weight and obese never smokers and then normal weight and obese ever-smokers, respectively. The mortality rates begin with a *k* and are labeled according to their corresponding compartment. For instance, *k_nwd_* is the mortality rate for normal weight diabetic never-smokers. Similarly, *k_nwds_* is the mortality rate for normal weight diabetic ever-smokers and is calculated as the baseline mortality rate plus the smoking mortality rate add-on value. All other parameters are transition rates between disease states.

**Table A2:** Transition Rate Parameter Names

| Parameters description | Parameter name | Models |
| --- | --- | --- |
| Normal weight never-smoking to obese never-smoking | $k_1$ | All models |
| Obese never-smoking to normal weight never-smoking | $k_2$ | All models |
| Smoking initiation rate | $k_3$ | All models |
| Normal weight ever-smoking to obese ever-smoking | $k_4$ | All models |
| Obese ever-smoking to normal weight ever-smoking | $k_5$ | All models |
| COPD incidence rate | $k_{10}$ | Model 3 and combined model |
| Normal weight ever-smoking with COPD to obese ever-smoking with COPD | $k_{11}$ | Model 3 and combined model |
| Obese ever-smoking with COPD to normal weight ever-smoking with COPD | $k_{12}$ | Model 3 and combined model |
| Cachexia initiation rate | $k_{13}$ | Model 3 and combined model |

### A1.6 Simplified Model 1 and 2 Equations

Below are the ODEs used to simulate the flows between disease states for the simplified model that includes state transitions for diabetic individuals only. There is only one set of equations for Model 1 while each age group has its own set of equations, transition and mortality rates in Model 2. The model schematic is shown in Figure 2(b). The model equations are given by:

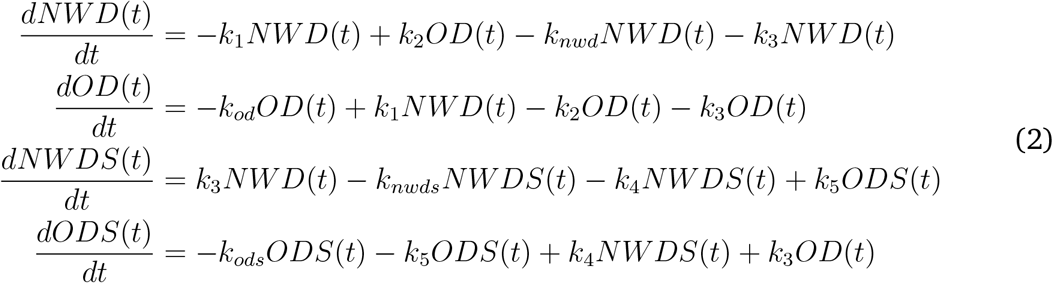

where *NWD* and *OD* are normal weight and obese diabetic never-smokers, respectively, and *NWDS* and *ODS* are the corresponding normal weight and obese ever-smokers. *k*_1_ is the rate at which normal weight never-smoking individuals become obese. *k*_2_ is the rate at which obese never-smoking individuals become normal weight. *k*_3_ is the smoking initiation rate. *k*_4_ is the rate at which normal weight ever-smoking individuals become obese. *k*_5_ is the rate at which obese ever-smoking individuals become normal weight. The mortality rates begin with a *k* and are labeled according to their corresponding compartment. For instance, *k_nwd_* is the mortality rate for normal weight diabetic never-smokers. All other parameters are transition rates between disease states. The initial state variables (initial conditions) are in units of people.

### A1.7 Model 2: Adding Age to the Original CM

Although age was not explicitly depicted on the original DAG (Figure 2(a)), the analysis conducted by Preston et al. standardized mortality rates according to US census ages. Because age is a confounder in the relationship between the exposure, BMI and outcome, mortality, we should adjust for it in the statistical analysis to obtain an unbiased effect estimate. However, we initially ran the same statistical analysis as done for model 1 to see if not adjusting for age correctly could result in the obesity paradox. The purpose of this exercise is analogous to a sensitivity analysis in that, we investigate how unmeasured bias may have altered our study data and results.

To incorporate this into our study, we split our population into a younger age-group (ages 40-59) and an older age-group (ages 60-74) to explore how age-varying rates might lead to the obesity paradox. Among older adults (i.e., our study population), age affects obesity status [52], diabetes status [53], and mortality. Smoking initiation rates are quite low after age 40 i.e., ~1% so we will assume that this rate is the same regardless of age-group [54]. See Figure 2(c) for the Model 2 DAG.

Apart from considering age, The model schematic (Figure 2(b)) and model equations for Model 2 are the same as Model 1 (Equations (2)). To add in age-varying rates, we included one set of equations for the younger age group and one identical set of equations for the older age group. The equations are otherwise the same, but parameters and initial conditions vary between age-groups, which accounts for the effects of age. We assumed that everyone remains in their given age group over the course of the study (one year).

### A1.8 Age-Weighting for Age-Structured Models

We age-weighted our model using weights from the 2010 census [19]. Specifically, the proportion of individuals in the young age group (ages 40-59) in the US population is 0.2771295 while the proportion of individuals in old age group (ages 60-74) is 0.1247997. Thus for a total study population of 1,000,000 individuals, there are

- 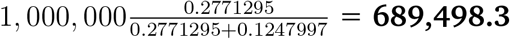 young individuals
- 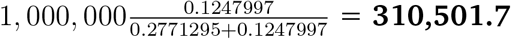 old individuals

### A1.9 Model 3: Adding Reverse Causation in the Original Model

We next tested the hypothesis that reverse causation may cause the obesity paradox. We first assumed that our observational study design was the same as in previous models and that we did not account for reverse causation in our data collection or statistical analyses (i.e., we ran the statistical analysis based on the original DAG in Figure 2(a)). We made changes directly to the model to test this alternative underlying causal mechanism and made a new corresponding DAG. See Figures 2f and 2e for the new model and corresponding DAG, and see Appendix Equations (3) for the model equations.

As mentioned previously, complications from comorbid diabetes and other diseases such as COPD may induce weight loss [20, 21] and also increase the risk of mortality [22, 23]. We therefore extended Model 1 to simulate how undiagnosed COPD and associated complications may be a risk factor for mortality and also affect the exposure, BMI. Because these complications affect the outcome and exposure, this model incorporates reverse causation in that the higher risk of mortality may precede changes in the exposure. In our extended model, normal weight and obese ever-smokers can transition into COPD disease states, marked with a ‘*C*’. We assumed that comorbidity of diabetes and COPD only occurs among ever-smokers since smoking is a key risk factor for COPD [55]. Additionally in our extended model, individuals with comorbid diabetes and COPD can then transition into the ‘unhealthy’ compartment, *U*. Individuals in *U* have lost weight due to cachexia and also have higher mortality rates than their normal weight ‘healthy’ counterparts (i.e. normal weight ever-smoking individuals with COPD who have not undergone cachexia). We assumed that BMI does not affect the rate at which individuals get COPD or transition into *U*. In some cases (depending on parameter values), individuals in *U* may also have higher mortality rates than obese ever-smoking individuals with COPD. Importantly, for the statistical analysis, ‘unhealthy’ individuals are measured as normal weight ever-smoking diabetics since our original study design did not measure COPD or the occurrence of cachexia. Individuals with diabetes are at an increased risk for developing COPD [56], so it is also possible that even if individuals with COPD at baseline in our cohort study were excluded, participants may have developed COPD and moved into the unhealthy disease state over the course of the study (this would be more likely for longer prospective studies more than 1 year). We examined this in two sensitivity analyses in which we exclude individuals with COPD at baseline and run the study for 1 year and 5 years.

Because this reverse causation mechanism relies on exposure status changing due to a risk factor for the outcome, mortality, the corresponding DAG is longitudinal to represent time-varying exposure and covariates. Specifically, this DAG incorporates changes to BMI over time. The exposure is *BMI*_0_ the BMI measurement at baseline, and the outcome is cumulative mortality at the end of the study. More details for converting between longitudinal DAGs and CMs can be found in [1].

We note that in this case when mortality is the outcome reverse causation can be described as confounding by disease (i.e., disease affects both weight loss and mortality) [24]. However, to be consistent with the nomenclature of Preston et al. [9] and others, we will refer to this mechanism as reverse causation.

### A1.10 Reverse causation Model 3 Equations

Below are the ODEs used to simulate the flows between disease states for the reverse causation model. The model schematic is shown in Figure 2(f). The model equations are given by:

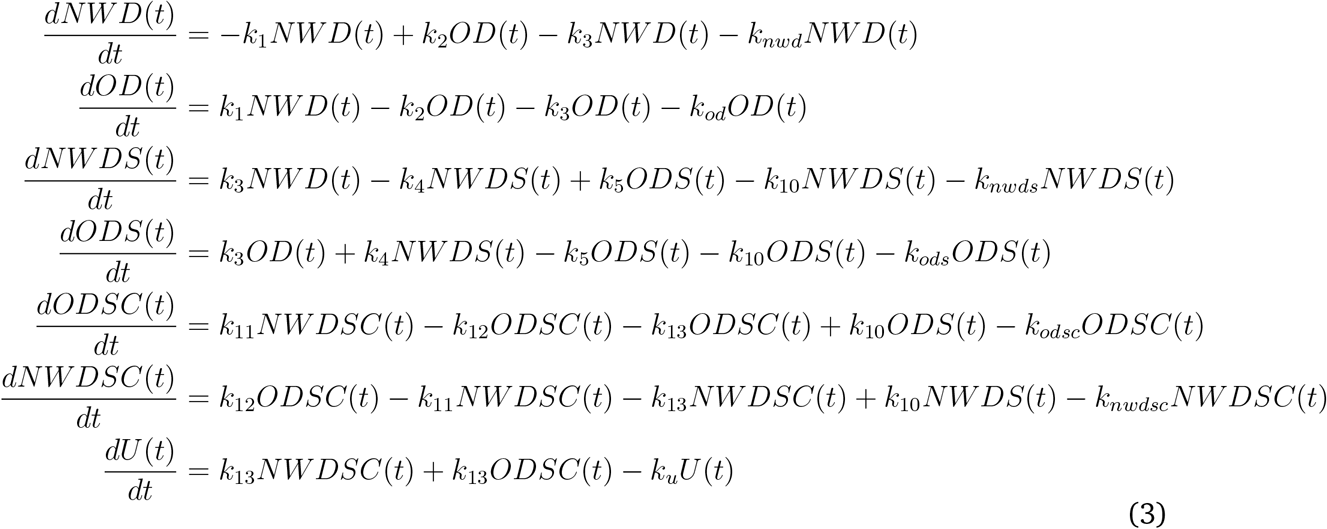

where *NWD* represents normal weight diabetic individuals; *OD* represents obese diabetic individuals; *ODS* are obese diabetic ever-smokers; *NWDS* are normal weight diabetic ever-smokers; *ODSC* and *NWDSC* are obese and normal weight ever-smoking diabetic individuals with COPD; and *U* are unhealthy individuals with comorbid COPD and diabetes who have undergone cachexia. These individuals have higher mortality rates than their unhealthy counterparts (i.e., *NWDSC*) and in some cases than *ODSC* but are measured together with healthy normal weight individuals due to the design of our observational study. *k*_1_ is the rate at which normal weight never-smoking individuals become obese. *k*_2_ is the rate at which obese never-smoking individuals become normal weight. *k*_3_ is the smoking initiation rate. *k*_4_ is the rate at which normal weight ever-smoking individuals become obese. *k*_5_ is the rate at which obese ever-smoking individuals become normal weight. *k*_10_ is the COPD incidence rate. *k*_11_ is the rate at which normal weight individuals with COPD become obese. *k*_12_ is the rate at which obese individuals with COPD become normal weight. Finally, *k*_13_ is the cachexia incidence rate. Finally, mortality rates are labeled according to their corresponding compartment. For instance, *k_nwds_* is the mortality rate for normal weight diabetic ever-smokers and is calculated as the baseline mortality rate plus the smoking mortality rate add-on value. All other parameters are transition rates between disease states. The initial state variables (initial conditions) are in units of people.

### A1.11 Combined Model

In the combined model, we incorporated both reverse causation and age-dependant mortality. Apart from considering age, the schematic (Figure 2(f)) and ODE equations (3) are the same as Model 3, but are vectorized such that each age group has its own set of equations. See Figure 2(g) for corresponding DAG which incorporates both reverse causation and age varying mortality. Because COPD prevalence increases with age [57], we allowed rates of transition to COPD to vary between age groups. Furthermore, because cachexia increases with age [58], we allowed rates of transition to *U* to vary between age groups. Finally, as reflected in Model 2, age affects BMI, diabetes and mortality. The MRR calculations are the same as conducted for the other models. We also ran various sensitivity analyses to see if adjusting for bias can keep the obesity paradox from occurring. Specifically, we (1) adjusted for age by standardizing to the unexposed population, (2) excluded individuals with COPD at baseline, and (3) combined 1 and 2.

### A1.12 Initial Condition Calculations

To determine how to distribute the population across disease states, the proportion of individuals in each state was randomly sampled using LHS [18]. For each model run, the sum of all sampled population fractions for the initial states must equal 1 to ensure uniformly sampled proportions. We randomly sampled proportions of the population in each disease state then multiplied the sampled proportions by the number of individuals in the given age group to get numbers of people starting in each disease state.

For instance in model 1 there are 4 states, we sampled 3 (total states - 1) values between [0,1]. Then we appended 0 and 1 onto the vector and sorted e.g., {0,0.1,0.4, 0.5, 1}, generating cut-points for the interval [0,1], allowing the interval to be divided uniformly at random among the four states. Next, we took the lagged differences between the elements in the vector i.e., in this example {0.1,0.3,0.1,0.5} to get the start proportions for each state. This process avoided sampling in a specific order (and leaving the last state to be determined by taking the 1 minus the previous), which would more frequently result in the last disease state have a lower proportion. We finally multiplied the proportions by census weights to determine how many individuals start in each state in this example: {40,192.92, 120,578.76, 40,192.92, 200,964.6}, totalling 401,929.2.

In our model, we intentionally did not use real data to inform our initial conditions, so that we could draw general qualitative conclusions about the structure of the model and its effects on our statistical results (i.e., our conclusions apply to a wide range of initial condition distributions). However, initial conditions can also be taken directly from the distribution of individuals across disease states in a study population. An alternative to this is starting with the distribution of individuals in a target population (i.e., the group of individuals that study results are applied to) and then simulating sampling procedures to explore how e.g., selection bias might affect the results. For instance, perhaps obese individuals from the younger age group are more likely to be included in the study than obese individuals from the older age group.

### A1.13 Mortality Rates

We imposed biologically realistic restrictions on the mortality rates such that ever-smokers have a higher mortality rate than their never-smoking counterparts (i.e., within weight strata), and obese individuals have a higher mortality rate than their normal weight counterparts i.e., within smoking strata).

Specifically, we set *ODS* mortality ≥ *NWDS* mortality ≥ *NWD* mortality and *ODS* mortality ≥ *OD* mortality ≥ *NWD* mortality. We did this by sampling a baseline mortality rate between 1% to 10% per year and then sampled ‘add-on’ mortality rates between 0% and 10% for obesity and separately smoking, thus the minimum mortality rate for obese, diabetic ever-smokers is 1% and the maximum is 30% per year. For instance, the mortality rate for smoking is the baseline mortality rate plus the smoking mortality rate add-on value.

#### A1.13.1 Model 2: Age-varying mortality

All transition rates between disease states were allowed to vary by age group with the exception of smoking initiation. We set smoking initiation to be the same, because as mentioned, we assumed that the initiation rates were quite low anyway in these ages i.e., after 40 years of age [54]. Older age group mortality rates for a given disease state were determined by multiplying the younger age group mortality rate of the same state by a scaling factor between 1 and 2. We chose a maximum of 2 because it is a rough approximation of the relative mortality rates for the younger compared to older age groups in the US according to the Centers for Disease Control and Prevention [59], although the age-groups are slightly different than in our model. Overall, within a given age-group the same restrictions on the relative mortality rates across disease states were used (as was used in Model 1). See Table 1 for all parameter ranges.

#### A1.13.2 Model 3: Reverse Causation

We placed the same biologically plausible restrictions on the relative mortality rates as we did for Model 1 (i.e., a baseline mortality rate and add-ons for obesity, and smoking) and included an additional add-on for COPD related mortality. Finally, we derived the mortality rate in the *U* compartment by multiplying the mortality rate of normal weight diabetic healthy ever-smokers with COPD by a cachexia scaling factor between 1 and 2 (similar to the age scaling factor in Model 2). We chose a maximum of 2 since it was a relatively conservative estimate that corresponded with the age-varying mortality rate. This enabled us to directly compare causal mechanisms without making assumptions about the relative rates for age compared with cachexia associated mortality. Thus, the maximum mortality rate of normal weight diabetic unhealthy ever-smokers was equal to 80%. See Table 1 for all parameter ranges.

#### A1.14 Person-Time for Simulated Studies

Even though the CM parameters were in units of years, the time steps for our model were in days. This is due to the fact that the sampling of both the initial conditions and parameter values could potentially lead to very fast, transient dynamics at the beginning of the simulation. For instance, if the transition rates out of the obese compartment are very fast and the initial conditions place the majority of individuals in the obese compartment, there will be a rapid decline in the numbers of obese individuals in the early stages of the simulation. This is not realistic especially in older age groups. Therefore, dividing our one-year time step into days enabled us to get a more precise estimate of the mean person-time spent in each compartment.

We calculated person-time by taking the daily average number of individuals in each disease state for the study. Specifically, we approximated the number of individuals in each state at each timestep using the life table method [60] (which is analogous to the trapezoidal rule) in which for a given timestep *t* the number of people in a state at time *t* and time *t* + 1 is averaged. See Appendix Equation (4) for details. We then added all person-days and converted to person-years by dividing the sum by 365 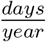 to get a final value in person-years. For slower dynamics or a longer run study, we could have calculated personA1.15 Trapezoidal Rule for Person-Time Calculation years without averaging over each day.

#### A1.15 Trapezoidal Rule for Person-Time Calculation

We calculated person time for a given time step, *t*, in simulations using the following equation i.e. the trapezoidal rule, equivalent to the life table method [60] in which all withdrawals or deaths are assumed to happen at the midpoint of each interval:

*Equation for Person Time*

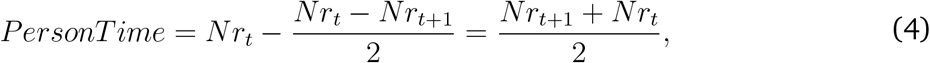

where *‘Nr_t_’* is number of individuals in a given compartment at time *t* and *’Nr*_t+1_’ is number of individuals in the same compartment at time *t* + 1.

#### A1.16 Incident Mortality for Simulated Studies

Next, we calculated incident mortality, the outcome, according to the following equation:

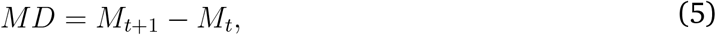

where *MD* is the incident mortality, *M_t_* is cumulative number of deaths in a given compartment at time *t* and *M_t+_*_1_ is the cumulative number of deaths in the same compartment at time *t* + 1. The cumulative number of deaths by compartment was quantified by adding extra equations with only the death rate for given compartment multiplied by the number of people in that compartment. We calculated the total incident mortality numbers for the entire year by each disease state. We next split our dataset into ever-smoking diabetics and never-smoking diabetics. We calculated a unique MRR for each strata. This accounts for the role of smoking as a confounder. See Appendix Section A1.18 for an example simulated dataset for a single study.

#### A1.17 Obesity Paradox Mortality Rate Parameterization

We can approximately back-calculate the mortality rate for a given CM compartment (i.e. the rate determined by LHS) by taking the total number of deaths over the course of simulation for that compartment (Equation (5)) divided by the person time approximation for that compartment (Equation (4)). For instance, in Table A3, the mortality rate of normal weight never-smokers used in the model is just 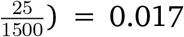 deaths per-year. We can also calculate the mortality rate ratio of normal weight compared to obese never-smokers by hand from the simulated dataset. For instance, in Table A3, we can just divide the mortality rate of normal weight individuals 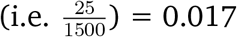 deaths per-year) by the mortality rate of obese individuals 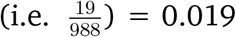 deaths per-year) to get an MRR of 0.895. The final MRRs obtained from our analysis are simply the ratio of CM mortality rates.

Alternatively, if we want to parameterize a CM from real-world data, we can use MRRs by taking the exponentiated beta estimates from a Poisson model. For instance, if the MRR of normal weight, never-smoking diabetics compared to obese never-smoking diabetics is 1.5 we know that the ratio of mortality rates among these two compartments is 1.5 (e.g. they could be 0.3 to 0.2). Now, if our model among never-smoking diabetics is the same as the crude Poisson model equation above (See equation below (6)), we can also take the exponentiated 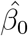 to obtain the mortality rate among normal weight, never-smoking diabetics and then use the MRR to determine the mortality rate among obese never-smoking diabetics.

#### A1.17.1 Poisson Model

Here, we show how to run a Poisson regression model from simulated data. This was not used in our analysis because we didn’t incorporate any type of sampling error into our model. However, one may want to simulate sampling using a multinomial draw in which case a Poisson regression model would be appropriate and could be used to derive confidence intervals to account for sampling error.

To run a standard Poisson regression model on our simulated dataset from Model 1 (see Table A3 for example data), we can calculate mortality rate ratios representing the effect of normal weight compared to obese individuals on mortality:

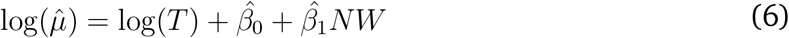

where log(*T*) is *log*(*person – time*) and is also the offset term which accounts for unequal follow up times between compartments and allows us to model the rate. The outcome, 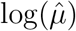, is the estimated incident mortality rate and 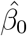 is log(incident mortality rate) among obese diabetics (i.e. when normal weight (*NW*) is equal to 0). Finally, 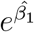 is the mortality rate ratio comparing mortality among normal weight individuals to mortality among obese individuals. It is also the multiplicative effect on the mortality rate of being obese compared to being normal weight.

Note that it is also possible to individuate our simulated population by sampling according to a standard population e.g. the census and then to run a different type of regression model for count data, e.g. Cox proportional hazards, using individual (not compartment) level data. However, the Poisson regression model is simpler to implement and an appropriate choice for count data which is a commonly assumed in epidemiological studies.

#### A1.18 Example Simulated Dataset

Table A3 shows an example dataset generated from the CM output among never-smokers. For each characteristic, ‘yes’ is coded as 1 and ‘no’ is coded as 0. The first row represents individuals in the obese diabetic never-smoking compartment while the second row represents individuals in the normal weight diabetic never-smoking compartment. The MRR comparing normal weight never-smokers to their obese counterparts for this given parameter set is therefore 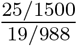.

**Table A3:**
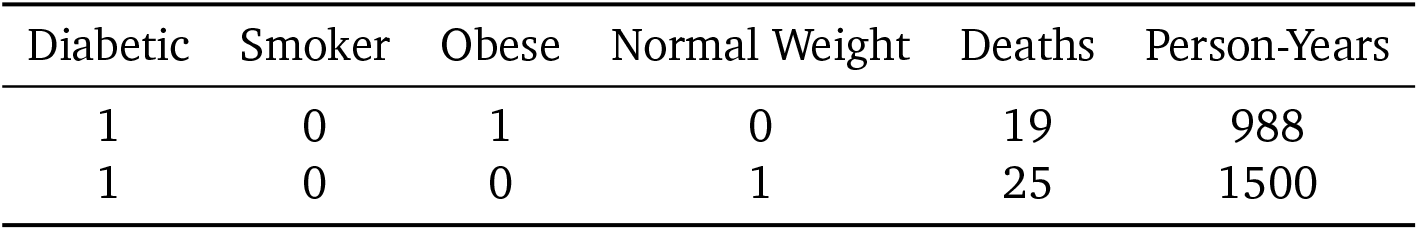
Example Simulated Dataset Among Never-Smokers

| Diabetic | Smoker | Obese | Normal Weight | Deaths | Person-Years |
| --- | --- | --- | --- | --- | --- |
| 1 | 0 | 1 | 0 | 19 | 988 |
| 1 | 0 | 0 | 1 | 25 | 1500 |

#### A1.19 Gamma Distributed Transition Rates

We incorporated an approximately gamma distributed transition rate from normal weight individuals with COPD to cachexia (accomplished by replacing the direct flow from COPD to cachexia with a sequence of compartments [42]) in the reverse causation model to see how relaxing our simplifying assumption of exponential transition rates might affect our conclusions. We found that incorporation of the gamma-distributed transition somewhat reduced the number parameter sets that resulted in the obesity paradox in the 5-year adjusted analysis (i.e., after excluding individuals with COPD at baseline and running the study for 5 years), but did not otherwise change the overall results (shown in Figure A4).

**Figure A3:**
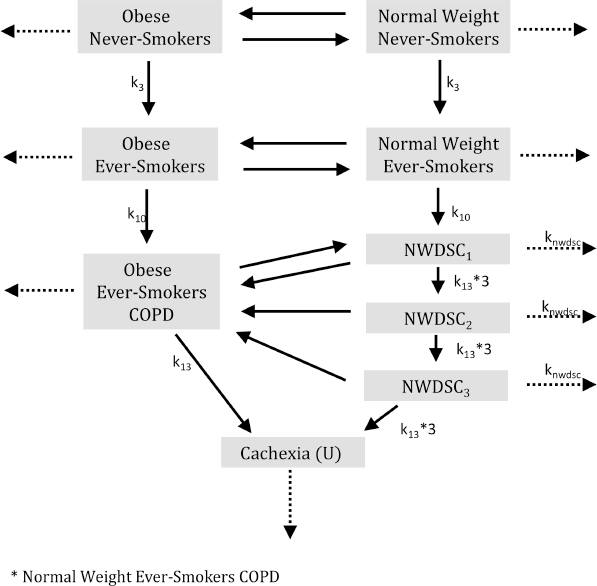
Schematic of reverse causation CM with gamma distributed transition rate between normal weight individuals with COPD to cachexia. For rate naming conventions, refer to Appendix Equations (3).

**Figure A4:**
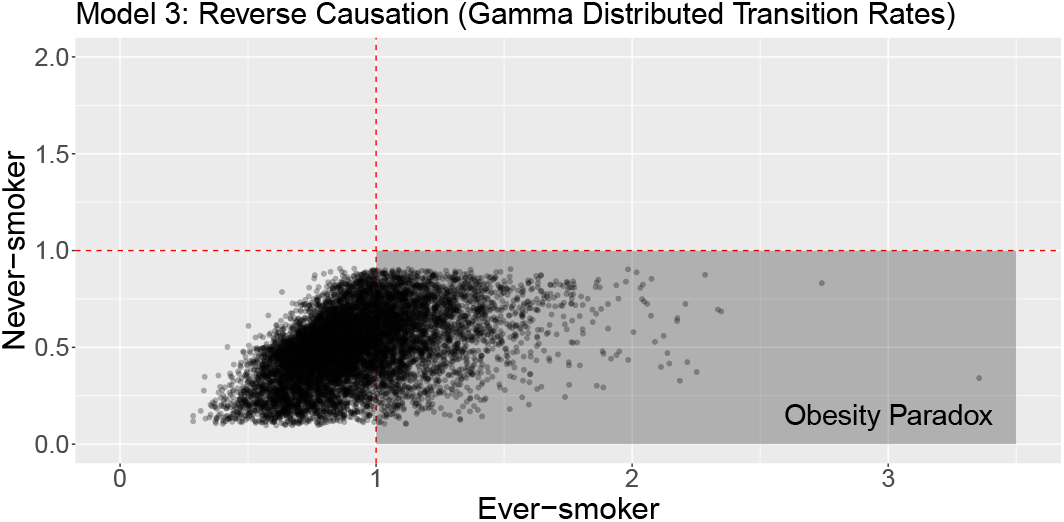
Results from the reverse causation CM with gamma distributed transition rate between normal weight individuals with COPD to cachexia. The plot shows the MRR (mortality rate ratio) comparing normal weight individuals to obese individuals for never-smokers against the corresponding MRR for ever-smokers for each of the 10,000 LH-sampled parameter sets with each point representing a single simulated study. The obesity paradox occurs when obese never-smoking diabetics have higher rates of mortality than normal weight never-smoking diabetics and obese ever-smoking diabetics have lower rates of mortality than normal weight ever-smoking diabetics.

## Footnotes

1 However, we note that DAGs technically also require parsimony in that (for example), all mediators between a given cause and effect are not included. Additionally, in practice DAGs often do not include all common causes, e.g. if it is not fully clear whether certain features are causally related.

## Notes

### Competing Interest Statement

The authors have declared no competing interest.

### Funding Statement

This study was funded by National Institute of General Medical Sciences Grant U01GM110712.

### Author Declarations

All relevant ethical guidelines have been followed and any necessary IRB and/or ethics committee approvals have been obtained.

Any clinical trials involved have been registered with an ICMJE-approved registry such as ClinicalTrials.gov and the trial ID is included in the manuscript.

